# The Longitudinal Aging Study Amsterdam COVID-19 exposure index: a cross-sectional analysis of the impact of the pandemic on daily functioning of older adults

**DOI:** 10.1101/2022.02.02.22270309

**Authors:** Emiel O. Hoogendijk, Noah A. Schuster, Theo G. van Tilburg, Laura A. Schaap, Bianca Suanet, Sascha de Breij, Almar A.L. Kok, Natasja M. van Schoor, Erik J. Timmermans, Renate T. de Jongh, Marjolein Visser, Martijn Huisman

## Abstract

**Objectives:** The aim of this study was to develop an index to measure older adults’ exposure to the COVID-19 pandemic, and to study its association with various domains of functioning.

**Design:** Cross-sectional study.

**Setting:** The Longitudinal Aging Study Amsterdam (LASA), a cohort study in the Netherlands.

**Participants:** Community dwelling older adults aged 62-102 years (n=1089) who participated in the LASA COVID-19 study (June-September 2020), just after the first wave of the pandemic.

**Primary outcome measures:** A 35-item COVID-19 exposure index with a score ranging between 0 and 1 was developed, including items that assess the extent to which the COVID-19 situation affected daily lives of older adults. Descriptive characteristics of the index were studied, stratified by several socio-demographic factors. Logistic regression analyses were performed to study associations between the exposure index and several indicators of functioning (functional limitations, anxiety, depression, and loneliness).

**Results:** The mean COVID-19 exposure index score was 0.20 (SD 0.10). Scores were relatively high among women and in the southern region of the Netherlands. In models adjusted for socio-demographic factors and pre-pandemic functioning (2018-2019), those with scores in the highest tertile of the exposure index were more likely to report functional limitations (OR: 2.23; 95% CI: 1.48 to 3.38), anxiety symptoms (OR: 3.87; 95% CI: 2.27 to 6.61), depressive symptoms (OR: 2.45; 95% CI: 1.53 to 3.93) and loneliness (OR: 2.97; 95% CI: 2.08-4.26) than those in the lowest tertile.

**Conclusions:** Among older adults in the Netherlands, those with higher scores on a COVID-19 exposure index reported worse functioning in the physical, mental and social domain. The index may be used to identify persons for whom targeted interventions are needed to maintain or improve functioning during the pandemic or post-pandemic.

**Strengths and limitations of this study:**

- This study was based on a representative sample of older adults from three culturally different regions in the Netherlands.
- The Longitudinal Aging Study Amsterdam COVID-19 study provides unique data on functioning of older adults in various life domains during the COVID-19 pandemic.
- The items of the COVID-19 exposure index that was developed were based on self-report, more objective sources such as medical records were lacking.
- The study covered the first months of the pandemic in the Netherlands, longitudinal data is needed to monitor functioning of older adults in later stages of the pandemic.

## Introduction

The COVID-19 pandemic has affected the daily lives of many older people, directly or indirectly. Some older adults or their close relatives have experienced the disease themselves, while others mainly experienced changes in their daily life related to measures taken by the government, such as lockdown and social distancing policies. Although the long-term effects of the pandemic on wellbeing and functioning of older adults are still unknown, there have been concerns about its negative effects on physical, psychological, and social functioning.^1–3^ Therefore, change in functioning of older adults during the pandemic has been the subject of various recent investigations.^1,4,5^ However, these studies seldom incorporate measures of actual exposure to pandemic-related events and situations.

Previous studies on the impact of the pandemic on functioning of older adults were predominantly conducted in the field of mental health, and show mixed results with regard to effects on psychological health outcomes, ^1,5–9^ both based on longitudinal^5,6^ or cross-sectional data.^1,7–9^ All studies have in common the assumption that the included individuals were equally exposed to the COVID-19 pandemic. However, the extent to which people have been exposed to COVID-19 and to related governmental measures varies widely within the older population. Older adults may, amongst others, have been confronted with infection and sickness, hospitalisation, loss of income, loss of contact with friends or family, infection and sickness of family members and even death of important others. Quantifying this exposure would help to identify persons that are most strongly affected by the pandemic, and would enable the monitoring of functioning of these people in the short- and long-term, to see whether tailored interventions are needed.

Measuring COVID-19 exposure can be achieved in various ways. So far, previous work focused on exposure to the actual COVID-19 infection only.^10,11^ Some studies used a broader definition, and explicitly asked about the impact of the pandemic on daily life.^12–15^ However, these studies were mostly focused on specific domains such as lifestyle,^14^ and were seldom conducted among older populations.^15^ Therefore, the aim of the current study was to develop an index to measure older adults’ exposure to the COVID-19 pandemic. Such an index summarizes the direct and indirect exposure to the pandemic on a broad range of topics relevant for older adults, such as COVID-19 infection and its consequences, financial problems, restrictions in healthcare use, social contact, and physical activity. Secondly, we aimed to study the associations of the index with various domains of functioning (i.e., physical, mental and social functioning). Data are used from the COVID-19 study that is part of the Longitudinal Aging Study Amsterdam (LASA), an ongoing population-based cohort study among older adults in the Netherlands.^16,17^

## Methods

### Study sample

LASA is an ongoing cohort study on various domains of functioning among older adults in the Netherlands.^16,18^ The study started in 1992, with follow-up observations approximately every three years. The study included older adults aged 55-84 years at baseline, based on a representative sample of the older population in three regions in the Netherlands. Refresher cohorts of older adults aged 55-64 years were added to the study in 2002 and 2012, using the same sampling frame. More details on the design, sampling and data collection of LASA have been reported elsewhere.^16,18,19^ In 2018-2019, the most recent regular LASA wave – with face-to-face and telephone interviews and clinical assessments – was completed, and the next wave was planned for 2021-2022. To monitor the impact of the COVID-19 pandemic more in-depth, an additional self-completion questionnaire was sent to participants on June 8, 2020, just after the first wave of the pandemic. The questionnaire included a broad range of measures to assess the impact of the COVID-19 situation on daily life, as well as a selection of measurements from regular LASA waves. In previous publications, the design and measurements of the LASA COVID-19 questionnaire were described in greater detail.^17,20,21^ The LASA study, including the COVID-19 study, received approval by the medical ethics committee of VU University medical centre. Written informed consent was provided by all respondents.

Eligibility criteria for the LASA COVID-19 study were: participation in the 2018-2019 wave (n = 1701) and being alive in March 2020 (n = 61 excluded). Furthermore, some respondents were excluded because filling out the questionnaire was expected to be too much of a burden for them. These were mainly people who had short or proxy interviews in the 2018-2019 wave (n = 155 excluded). This resulted in a selection of 1485 LASA respondents who received the COVID-19 questionnaire. Respondents were given the options to send back the questionnaire by mail or to fill out the questionnaire online (digital questionnaire). A telephone interview was offered to the oldest respondents (80+ years) who initially did not respond and for whom filling out a questionnaire appeared to be too difficult. Of 1485 respondents who received the questionnaire, 1128 (76%) participated. Data were received between June 9, 2020 and October 8, 2020. This included 909 written questionnaires, 198 digital questionnaires, and 21 telephone interviews. Of 1128 participants, 39 were excluded because of missing data on the newly developed COVID-19 exposure index, leaving an analytical sample of 1089 participants. For these participants, we also used in one of the analytical models (see below) pre-pandemic data on functioning from the 2018-2019 LASA wave.

### COVID-19 exposure index

The COVID-19 exposure index included variables that measured older adults’ direct and indirect exposure to the COVID-19 pandemic. This resulted in a 35-item index, including information on COVID-19 infection of respondents and their close relatives (including COVID-19-related hospitalisation and death), as well as items that assess the extent to which the COVID-19 situation affected healthcare use and access, providing and receiving personal care/homecare, the work situation, grocery shopping, lifestyle (e.g., physical activity and alcohol use), social behaviour, and various life events or situations, such as financial problems and leisure activities. For a full overview of all items, see Table 1. For the calculation of the index, we followed a method that is often used for calculating indexes in research among older populations, such as frailty indexes.^22,23^ For each item, scores were dichotomised as 0 (no) or 1 (yes). Subsequently, the exposure index score was calculated by dividing the sum of item scores present by the total number of item scores measured in a respondent (considering missing items when needed). This resulted in a score between 0 and 1, where higher scores indicate higher exposure to the COVID-19 pandemic. For example, in a person with nine positive items out of 35 measured items, the corresponding exposure index score is 9/35=0.26. We calculated the index score only if respondents were missing seven (20% of 35 items) or less item scores. Most older adults had no (73%) or one to three (22%) missing item scores.

**Table 1.** Overview of the variables included in the COVID-19 exposure index

| **Item** | **Cut-off** | **Prevalence (%)** |
| --- | --- | --- |
| 1. Tested positive for COVID-19 or probable COVID-19 (told by healthcare professional) | No = 0, Yes = 1 | 2.7 |
| 2. Hospital admission / ICU admission because of COVID-19 | No = 0, Yes = 1 | 0.4 |
| 3. Partner/parent/child with COVID-19 positive test | No = 0, Yes = 1 | 3.5 |
| 4. Partner/parent/child with COVID-19 hospital admission or death | No = 0, Yes = 1 | 1.3 |
| 5. Sibling/grandchild/other family member with COVID-19 hospital admission or death | No = 0, Yes = 1 | 5.3 |
| 6. Neighbour/friend/other acquaintance with COVID-19 hospital admission or death | No = 0, Yes = 1 | 30.0 |
| 7. Respondent has been in quarantine | No = 0, Yes = 1 | 11.8 |
| 8. GP visit cancelled by GP | No = 0, Yes = 1 | 9.5 |
| 9. GP visit replaced by telephone consultation | No = 0, Yes = 1 | 15.4 |
| 10. Respondent cancelled/postponed GP visit | No = 0, Yes = 1 | 6.9 |
| 11. Specialist outpatient visit cancelled by outpatient clinic | No = 0, Yes = 1 | 24.8 |
| 12. Specialist outpatient visit replaced by telephone consultation | No = 0, Yes = 1 | 21.4 |
| 13. Respondent cancelled/postponed specialist outpatient visit | No = 0, Yes = 1 | 7.5 |
| 14. Respondent postponed help seeking for physical/psychological complaints because of the COVID situation | No = 0, Yes = 1 | 8.3 |
| 15. Providing personal/household care: experience of increased burden during the COVID-19 pandemic | No = 0, Yes = 1 | 2.7 |
| 16. Providing personal/household care: more than before the COVID-19 pandemic | No = 0, Yes = 1 | 4.2 |
| 17. Decrease in received personal/household care during the COVID-19 pandemic | No = 0, Yes = 1 | 4.5 |
| 18. Work situation: lower salary due to the COVID-19 pandemic | No = 0, Yes = 1 | 1.0 |
| 19. Difficulties with grocery shopping during the COVID-19 pandemic | No = 0, Sometimes or always = 1 | 15.2 |
| 20. Weight loss / weight gain because of COVID-19 pandemic | No = 0, Sometimes or always = 1 | 37.7 |
| 21. Less physical activity than before the COVID-19 pandemic | No = 0, Sometimes or always = 1 | 49.8 |
| 22. Increased alcohol use during the COVID-19 pandemic | No = 0, Sometimes or always = 1 | 13.8 |
| 23. Less social contact with family during the COVID-19 pandemic | No = 0, Yes = 1 | 38.3 |
| 24. Less social contact with friends and acquaintances during the COVID-19 pandemic | No = 0, Yes = 1 | 40.9 |
| 25. Less social contact with formal relationships during the COVID-19 pandemic | No = 0, Yes = 1 | 12.3 |
| 26. Impact of job loss/financial problems of respondent during the COVID-19 pandemic | No impact = 0, Moderate or strong = 1 | 8.8 |
| 27. Impact of job loss/financial problems of close relative during the COVID-19 pandemic | No impact = 0, Moderate or strong = 1 | 15.5 |
| 28. Impact of cancelation of leisure activities during the COVID-19 pandemic | No impact = 0,<br>Moderate or strong<br>= 1 | 70.8 |
| 29. Impact of not being able to visit bars, restaurants and/or shops during the COVID-19 pandemic | No impact = 0,<br>Moderate or strong<br>= 1 | 72.5 |
| 30. Impact of experience of illness during the COVID-19 pandemic | No impact = 0,<br>Moderate or strong<br>= 1 | 10.5 |
| 31. Impact of death or severe illness of partner or household member during the COVID-19 pandemic | No impact = 0,<br>Moderate or strong<br>= 1 | 7.1 |
| 32. Death or severe illness of family member or friend during the COVID-19 pandemic | No impact = 0,<br>Moderate or strong<br>= 1 | 29.3 |
| 33. Impact of no contact or less contact with children/grandchildren during the COVID-19 pandemic | No impact = 0,<br>Moderate or strong<br>= 1 | 54.3 |
| 34. Impact of no contact or less contact with family/friends during the COVID-19 pandemic | No impact = 0,<br>Moderate or strong<br>= 1 | 67.3 |
| 35. Impact of difficulties in obtaining essential medication during the COVID-19 pandemic | No impact = 0,<br>Moderate or strong<br>= 1 | 5.8 |

### Outcomes

Various functional domains were evaluated. From the physical domain, functional limitations were assessed. Respondents were asked about any difficulty with performing seven basic activities of daily living: dressing and undressing, climbing the stairs, sitting down and getting up from a chair, cutting one’s own toenails, using transportation, walking five minutes outdoors without resting, and bathing. If respondents had difficulty with or could not perform at least one activity, functional limitations were considered to be present (no/yes). From the mental domain we included anxiety and depressive symptoms. Anxiety symptoms were measured using the anxiety subscale of the Hospital Anxiety and Depression Scale (HADS-A, range 0-21). A cut-off score of ≥8 was applied to indicate the presence of clinically significant anxiety symptoms.^24^ Depressive symptoms were assessed using the 10-item version of the Center for Epidemiologic Studies Depression Scale (CES-D-10, range 0-30). A cut-off score of ≥10 indicated the presence of clinically relevant depressive symptoms.^25^ Finally, the social domain was covered by loneliness, measured by the De Jong Gierveld Loneliness scale (range 0-11). We applied the cut-off score of ≥3 to indicate the presence of loneliness.^26^

### Covariates

Socio-demographic characteristics included sex, age, partner status, educational level, and region. Because of a non-linear association with most functional outcomes, age was categorised into: <70 years, 70-79 years, and ≥80 years. Partner status was defined as having a partner inside or outside the household (1) or having no partner (0). The highest level of completed education was assessed, and three categories were distinguished: low (elementary school or less), medium (lower vocational or general intermediate education), and high (intermediate vocational education, general secondary school, higher vocational education, college or university). A region variable indicated the three regions in which LASA respondents were recruited: the western part of the Netherlands (in and around Amsterdam), the northeast (in and around Zwolle), and the south (in and around Oss).

### Statistical analysis

First, descriptive statistics of the exposure index were calculated, such as mean, median and range. The distribution of the exposure index was presented with a histogram. Next, we described the demographic characteristics of the study population, for the total sample and stratified by categories of the exposure index. For the latter we divided the exposure index scores into tertiles, because there are no established cut-points. This approach is also helpful to gain insight into the potential dose-response relationship between the exposure index scores and various domains of functioning. Finally, we performed logistic regression analyses to study associations between the exposure index tertiles and functional limitations, anxiety, depression, and loneliness. Three models were fitted: a crude model, a model adjusted or age, sex, partner status, educational level and region, and a model additionally adjusted for pre-pandemic functioning (in 2018-2019). The latter was done to control for the possibility that people with worse pre-pandemic functioning had a higher likelihood of experiencing COVID-19 adversity (i.e., higher exposure index scores). In the 2018-2019 LASA wave, all functional indicators were defined in the same way and assessed with the same instruments as in the LASA COVID-19 study, as described above. All analyses were done in SPSS 26 (IBM Corp, Armonk, NY, USA).

## Results

In the current sample, the distribution of the COVID-19 exposure index was slightly skewed to the right (Figure 1). The mean exposure index score was 0.20 (SD 0.103), with a range from 0 to 0.63. The median was 0.20 (IQR: 0.13 to 0.27). Table 1 shows the prevalence of the items included in the exposure index, ranging from <1% (hospital admission or ICU admission of respondent because of COVID-19) to 73% (respondent experienced moderate or strong impact of not being able to visit bars, restaurants and/or shops during the COVID-19 pandemic).

**Figure 1.**
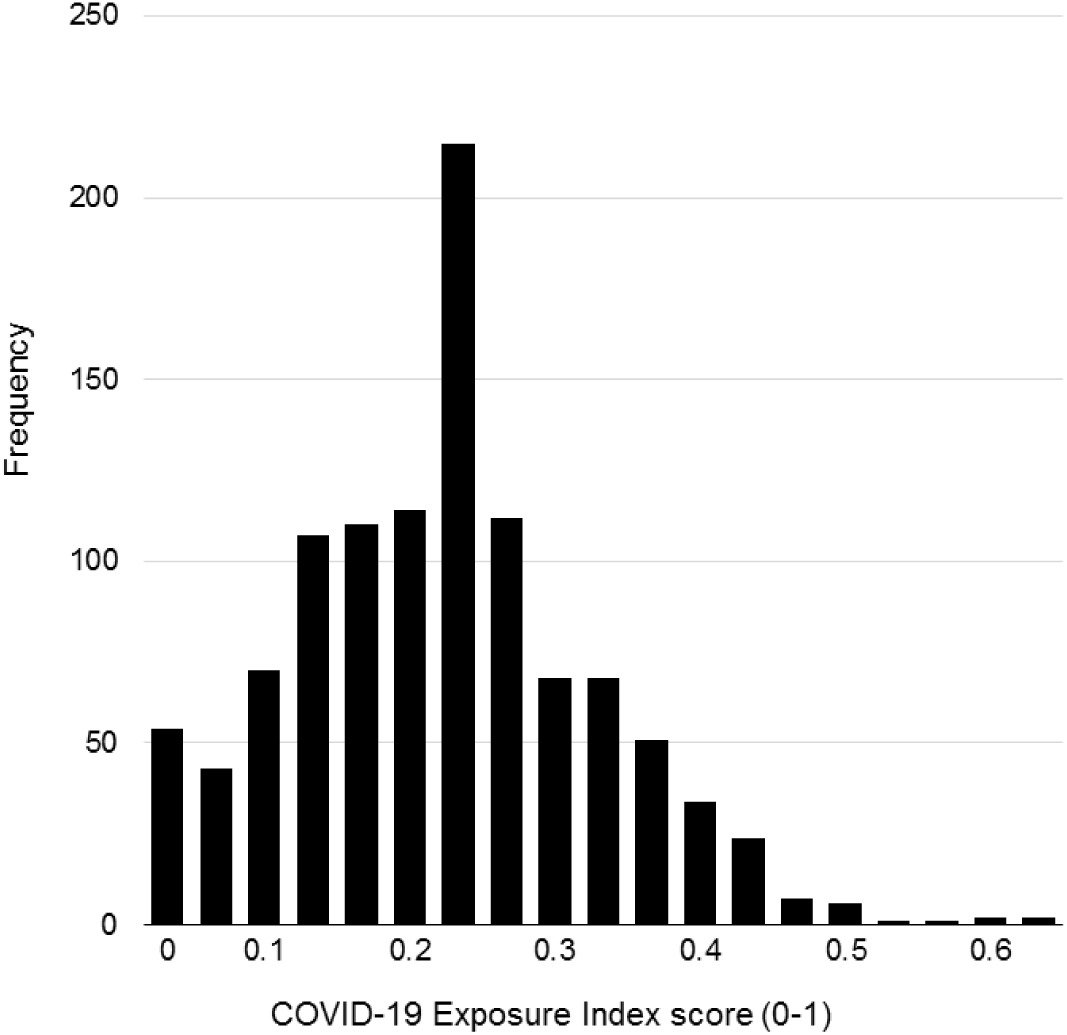
Distribution of the COVID-19 Exposure Index

The characteristics of the study sample are displayed in Table 2. The majority of the sample was female (53%), with partner (74%) and higher educated (55%). The largest proportion of the sample was aged between 70-79 years (43.8%) and lived in the Amsterdam region (43.7%). Table 2 also shows the characteristics of the study sample stratified by tertiles of the exposure index. COVID-19 exposure index scores were higher among women (57.3% of highest tertile vs. 43.4% of lowest tertile) and in the southern region of the Netherlands (29.6% of highest tertile vs. 19.5% of lowest tertile). No statistically significant differences in exposure index scores were observed for age categories, partner status and educational level.

**Table 2.** Socio-demographic characteristics of the total sample (n = 1089) and by COVID-19 exposure index tertiles

|  |  | Total | COVID-19 exposure index |  |  |  |
| --- | --- | --- | --- | --- | --- | --- |
|  |  |  | Lowest tertile | Middle tertile | Highest tertile |  |
|  |  |  | 0-0.142 | 0.143-0.235 | >0.235 |  |
|  |  | n (%) | n (%) | n (%) | n (%) | p |
| Sex | Men | 516 (47.4) | 209 (56.6) | 154 (42.5) | 153 (42.7) | <0.001 |
|  | Women | 573 (52.6) | 160 (43.4) | 208 (57.5) | 205 (57.3) |  |
| Age | <70 years | 384 (35.3) | 128 (34.7) | 128 (35.4) | 128 (35.8) | 0.95 |
|  | 70-79 years | 477 (43.8) | 166 (45.0) | 160 (44.2) | 151 (42.2) |  |
|  | ≥80 years | 228 (20.9) | 75 (20.3) | 74 (20.4) | 79 (22.1) |  |
| Partner status | With partner | 808 (74.2) | 287 (77.8) | 262 (72.4) | 259 (72.3) | 0.15 |
|  | No partner | 281 (25.8) | 82 (22.2) | 100 (27.6) | 99 (27.7) |  |
| Educational level | Low | 124 (11.4) | 49 (13.3) | 32 (8.8) | 43 (12.0) | 0.21 |
|  | Medium | 368 (33.8) | 128 (34.7) | 130 (35.9) | 110 (30.7) |  |
|  | High | 597 (54.8) | 192 (52.0) | 200 (55.2) | 205 (57.3) |  |
| Region | West (Amsterdam and surrounding) | 476 (43.7) | 180 (48.8) | 160 (44.2) | 136 (38.0) | <0.01 |
|  | Northeast (Zwolle and surrounding) | 359 (33.0) | 117 (31.7) | 126 (34.8) | 116 (32.4) |  |
|  | South (Oss and surrounding) | 254 (23.3) | 72 (19.5) | 76 (21.0) | 106 (29.6) |  |

P-values are based on Chi-square tests

Figure 2 shows the prevalence of functional limitations, anxiety symptoms, depressive symptoms and loneliness by tertiles of the COVID-19 exposure index. For all indicators, functioning was worse in those with scores in the highest tertiles of the exposure index. This was further confirmed in logistic regression analyses, in both crude and adjusted models (Table 3). In models adjusted for age, sex, partner status, educational level and region, people in the middle and highest tertile of the exposure index were more likely to have functional limitations (OR middle tertile: 1.61, 95% CI 1.15 to 2.26; OR highest tertile: 3.15, 95% CI 2.23 to 4.43) and loneliness (OR middle tertile: 1.84, 95% CI 1.35 to 2.50; OR highest tertile: 2.64, 95% CI 1.93 to 3.60) compared to people in the lowest tertile. For anxiety symptoms and depressive symptoms, only those in the highest tertile had a significantly higher probability of the outcome compared to the lowest tertile (anxiety symptoms OR: 3.84, 95% CI 2.25 to 6.65; depressive symptoms OR: 3.27, 95% CI 2.14 to 5.00). Further adjusting for pre-pandemic levels of functioning in the final model did not change the results: people in the highest tertile of the exposure index were more likely to report functional limitations (OR: 2.23; 95% CI: 1.48 to 3.38), anxiety symptoms (OR: 3.87; 95% CI: 2.27 to 6.61), depressive symptoms (OR: 2.45; 95% CI: 1.53 to 3.93) and loneliness (OR: 2.97; 95% CI: 2.08-4.26) than those in the lowest tertile.

**Table 3.** Logistic regression analyses: associations between the COVID-19 exposure index and various domains of functioning

|  | **Functional limitations** |  | **Anxiety symptoms** |  | **Depressive symptoms** |  | **Loneliness** |  |
| --- | --- | --- | --- | --- | --- | --- | --- | --- |
|  | n/N | OR (95% CI) | n/N | OR (95% CI) | n/N | OR (95% CI) | n/N | OR (95% CI) |
| **Crude model** |  |  |  |  |  |  |  |  |
| COVID-19 exposure index |  |  |  |  |  |  |  |  |
| Lowest tertile | 125/359 | 1.00 (ref.) | 20/361 | 1.00 (ref.) | 38/362 | 1.00 (ref.) | 136/366 | 1.00 (ref.) |
| Middle tertile | 159/361 | 1.47 (1.09-1.94) | 26/355 | 1.35 (0.74-2.46) | 57/356 | 1.63 (1.05-2.52) | 181/357 | 1.74 (1.29-2.34) |
| Highest tertile | 206/353 | 2.62 (1.94-3.55) | 66/351 | 3.95 (2.34-6.67) | 98/352 | 3.29 (2.19-4.95) | 212/357 | 2.47 (1.83-3.34) |
| **Adjusted for covariates\*** |  |  |  |  |  |  |  |  |
| COVID-19 exposure index |  |  |  |  |  |  |  |  |
| Lowest tertile | 125/359 | 1.00 (ref.) | 20/361 | 1.00 (ref.) | 38/362 | 1.00 (ref.) | 136/366 | 1.00 (ref.) |
| Middle tertile | 159/361 | 1.61 (1.15-2.26) | 26/355 | 1.31 (0.71-2.41) | 57/356 | 1.57 (1.00-2.46) | 181/357 | 1.84 (1.35-2.50) |
| Highest tertile | 206/353 | 3.15 (2.23-4.43) | 66/351 | 3.84 (2.25-6.55) | 98/352 | 3.27 (2.14-5.00) | 212/357 | 2.64 (1.93-3.60) |
| **Adjusted for pre-pandemic functioning\*\*** |  |  |  |  |  |  |  |  |
| COVID-19 exposure index |  |  |  |  |  |  |  |  |
| Lowest tertile | 115/325 | 1.00 (ref.) | 20/329 | 1.00 (ref.) | 35/330 | 1.00 (ref.) | 124/333 | 1.00 (ref.) |
| Middle tertile | 146/333 | 1.28 (0.85-1.93) | 24/328 | 1.33 (0.72-2.44) | 52/329 | 1.63 (0.99-2.67) | 166/329 | 1.94 (1.36-2.77) |
| Highest tertile | 190/326 | 2.23 (1.48-3.38) | 60/324 | 3.87 (2.27-6.61) | 86/325 | 2.45 (1.53-3.93) | 196/330 | 2.97 (2.08-4.26) |

\*adjusted for sex, age, partner status, educational level and region; \*\* additionally adjusted for pre-pandemic (2018-2019) functioning; n/N = number of positive outcomes/total N of the COVID-19 index tertile; OR = Odds Ratio; 95% CI = 95% Confidence Interval; ref. = reference category; Functional limitations = mild or severe limitations (yes); Anxiety symptoms = HADS-A $\geq 8$ ; Depressive symptoms = CES-D-10 $\geq 10$ ; Loneliness = De Jong Gierveld loneliness scale $\geq 3$ .

**Figure 2.**
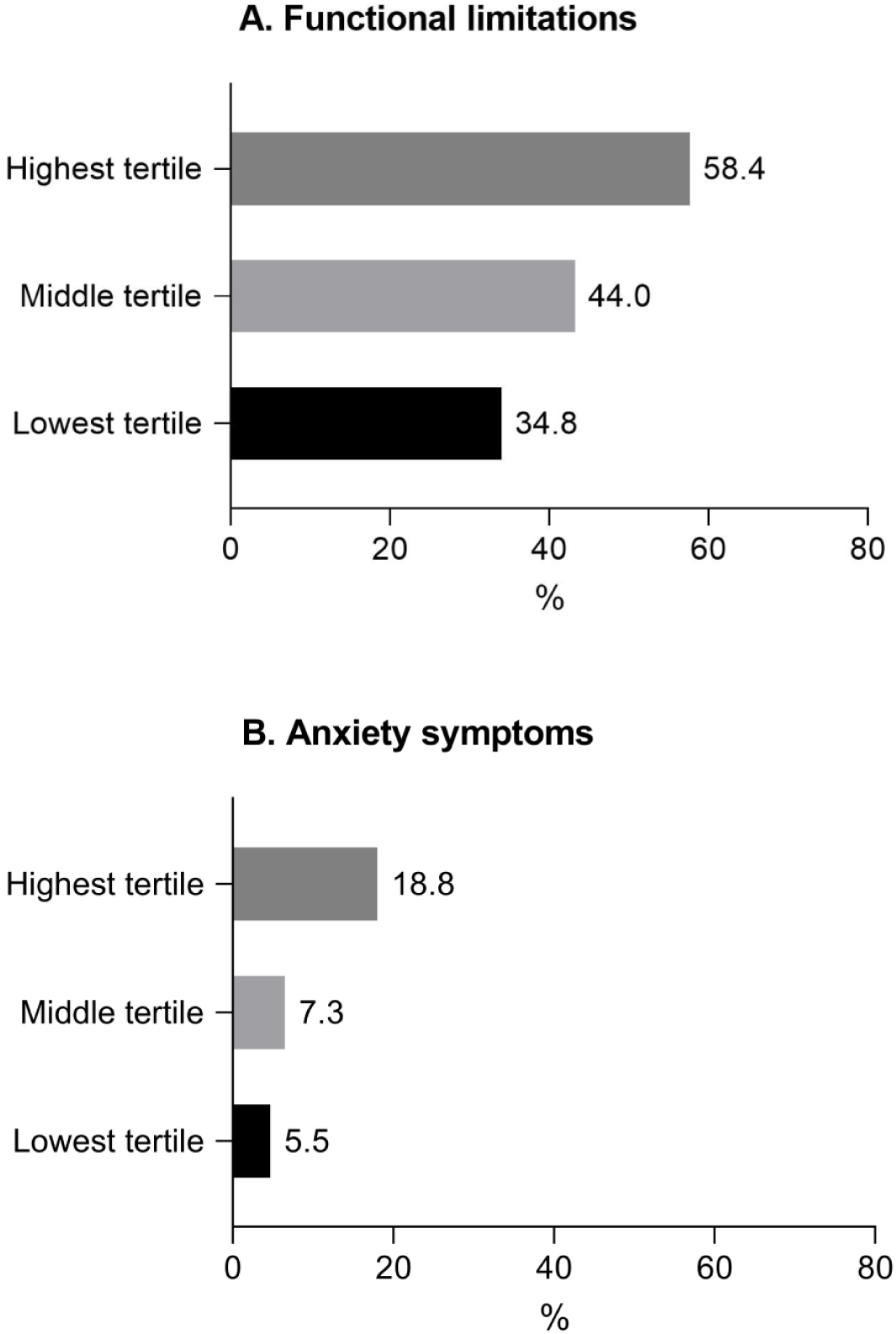
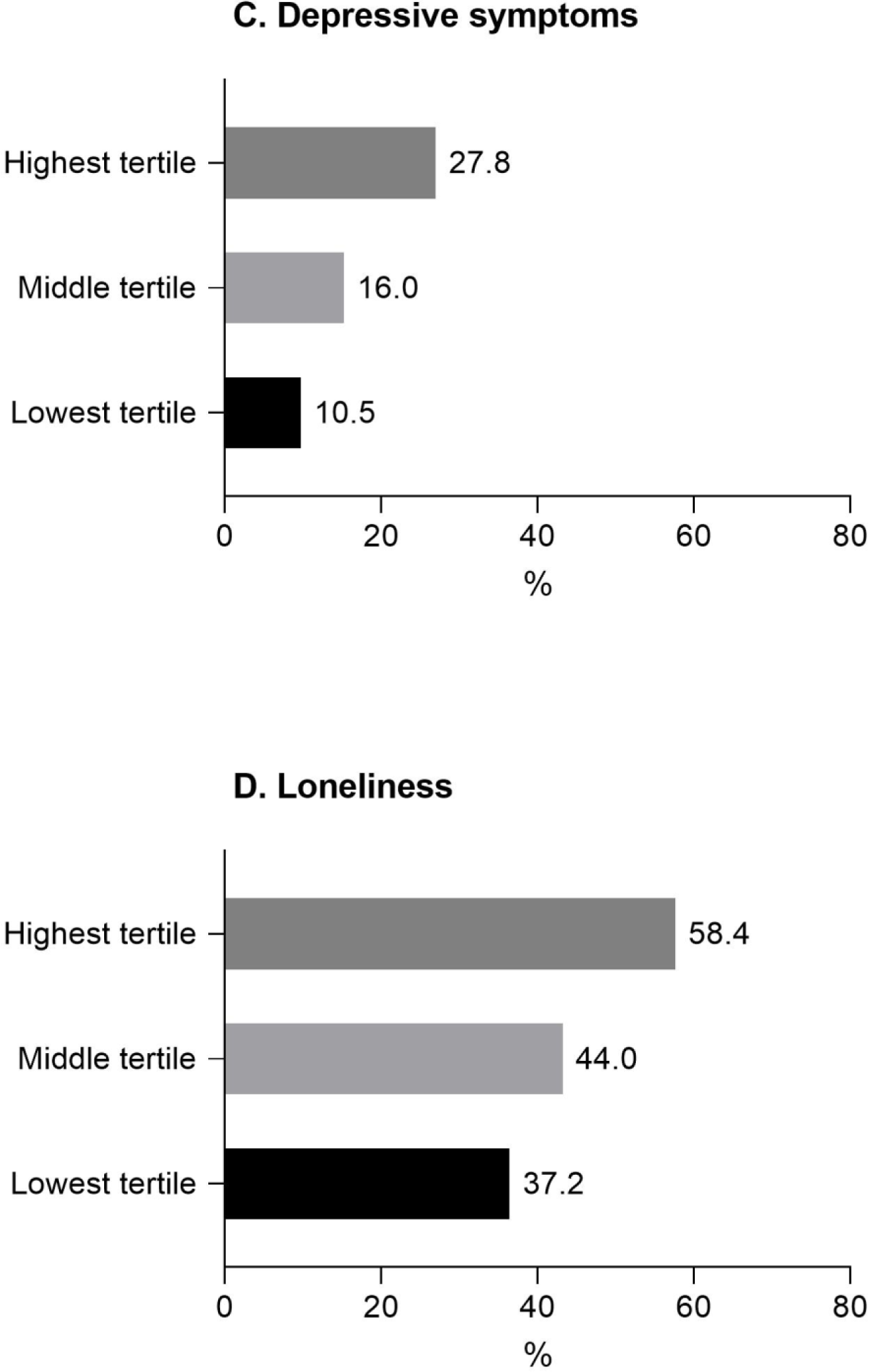
Various domains of functioning by COVID-19 exposure index tertiles: functional limitations (panel A), anxiety symptoms (panel B), depressive symptoms (panel C) and loneliness (panel D) Functional limitations = mild or severe limitations (yes); Anxiety symptoms = HADS-A ≥8; Depressive symptoms = CES-D-10 ≥10; Loneliness = De Jong Gierveld loneliness scale ≥3.

## Discussion

Using data from the LASA COVID-19 study in the Netherlands, collected just after the first wave of the pandemic, we developed an index to measure older adults’ exposure to the COVID-19 pandemic, and we studied associations of this index with various indicators of functioning. Our results revealed that older people with higher exposure index scores showed worse functioning across various domains, even after adjustment for pre-pandemic levels of functioning. This was observed in physical, mental and social domains, suggesting that the groups who experienced the greatest consequences from the COVID-19 situation also experienced relatively poor health and wellbeing across the board.

One previous publication described the development of a questionnaire to measure the impact of the COVID-19 pandemic on daily lives of older adults in the US.^15^ However, this publication does not contain any data. Therefore, our study is unique, and the findings cannot directly be compared to previous work. To determine whether exposure index scores are low or high, a comparison with older adults in other countries or at later time points during the pandemic is necessary. We have studied variation within our study sample and did not observe differences in exposure index scores by age and educational level. This indicates that during the first wave of the pandemic different demographic groups in the Dutch older population experienced a similar impact of COVID-19 and related governmental measures in daily life. In previous studies, socioeconomic differences in COVID-19-related morbidity and mortality have been reported in several countries.^27,28^ Our exposure index is a rather broad measure, not only covering COVID-19 related morbidity and mortality, which might explain the absence of an association with educational level. However, we did observe differences in exposure index scores by sex and region. Women are usually more socially active and provide more often informal care than men, and may have experienced stronger effects of governmental social distancing measures. The higher scores among LASA respondents in the south of the Netherlands was expected, since the southern regions were the epicentre of the Dutch COVID-19 outbreak in 2020.^17^

The findings of this study have practical implications, amongst others for health policy makers. It is of utmost importance to know to what extent the COVID-19 pandemic and related governmental measures affect daily functioning of older adults, positively or negatively. However, it is also important to take into account that there might be great variation within the older population with regard to experienced impact of the pandemic. The exposure index as created in the current study helps to capture this variation and may especially be useful to identify persons for whom targeted interventions are needed to improve or maintain functioning across various domains during the pandemic. This will contribute to improved health and wellbeing of older adults and may also help to develop health policy responses in future pandemics. For example, to prevent adverse effects on social functioning, strategies may be developed to maintain social contact during a pandemic.^29^

This was – to our knowledge – the first study to comprehensively measure the exposure to the COVID-19 pandemic among older adults. Where previous studies focused on isolated indicators of exposure such as infections,^10–13^ we created a more elaborated measure which summarised the consequences of the pandemic on everyday life. Our approach reveals that great heterogeneity exists within the older population in terms of impact of the pandemic on levels of functioning. Other strengths of the current study include the use of data from a large sample of older adults in the Netherlands, with indicators of functioning that covered multiple domains. However, the study also has limitations. First, our results should be interpreted with caution. Because of the cross-sectional design of the study, we have to be careful with drawing conclusions on the direction of the observed associations. We partly addressed this by controlling the analyses on the associations between the COVID-19 exposure index and various indicators of functioning for pre-pandemic functioning. This adjustment was needed, because it is possible that people who already had health problems before the pandemic have a higher chance to experience changes in healthcare or homecare, and therefore score higher on the COVID-19 exposure index 9. However, it is still possible that levels of functioning partly determine how people respond to certain items included in the exposure index. For example, mental health problems may result in a more negative evaluation of the impact of the pandemic on daily life. Second, our data covered the first wave of the pandemic in the Netherlands. We do not know to what extent these findings are generalizable to later stages of the pandemic and to other geographical areas. This will become more clear when follow-up data from the LASA study become available, as well as data from cohort studies in older populations across Europe, such as the SHARE study.^30,31^ Third, we created the exposure index with an existing dataset, so we were limited to variables available in this dataset. We missed consequences of the pandemic in some domains, such as concerns regarding the COVID-19 situation, the quality of sleep, and mood.^15^ Fourth, all the measures included in this study are based on self-report. For some items included in the exposure index, additional information from more objective sources such as medical records would have been helpful, especially with regard to details on COVID-19 infection, symptoms and test results. Finally, although the participation rate of the LASA COVID-19 study was rather high, we might have missed people in our sample with more severe COVID-19 infections. Therefore, the prevalence of some items in the exposure index could be an underestimation, such as COVID-19 infection and related hospitalisation.

There is a growing body of research exploring the effects of COVID-19 infection on morbidity and mortality in older adults.^32–35^ An increasing number of studies also has been investigating the impact of the COVID-19 pandemic on older adults’ levels of functioning in daily life, such as mental health, loneliness, lifestyle, and wellbeing.^5,21,36^ Yet, it is still unknown what the effects of the pandemic will be in the long-term. When monitoring levels of functioning of older adults over time and comparing them to pre-pandemic functioning, it is challenging to disentangle aging effects from changes that are due to the pandemic, which are period effects. The index that was developed in the current study could make this easier, because it enables the investigation of associations between COVID-19 exposure and long-term functioning. Therefore, an important direction of future research is to study associations between COVID-19 exposure and levels of functioning using long-term follow-up. Another question that remains unanswered, and that could be addressed in future research, is whether the COVID-19 exposure index itself changes over time. By repeatedly measuring the exposure index, patterns in COVID-19 exposure and the impact of cumulative exposure may be revealed. For example, it is possible that during the pandemic - in certain domains (e.g., healthcare provision, lifestyle or the work situation) - the impact of COVID-19 decreases due to adaptation processes, or that people feel less restricted and more safe because of increasing vaccination rates.

## Conclusion

We developed a COVID-19 exposure index using data from older adults participating in the LASA study in the Netherlands. We found that, just after the first wave of the pandemic, exposure was relatively higher among women and in the southern region of the Netherlands. Moreover, older adults with higher scores on the index reported worse functioning in the physical, mental and social domain. Our index may provide a more comprehensive and sensitive measure of COVID-19 exposure than measuring exposure based on infection alone. When monitoring functioning of older adults over time, the use of indexes such as ours enables the identification of people for whom targeted interventions are needed to maintain or improve functioning across various domains, during the pandemic or post-pandemic.

## Data Availability

All data underlying the results presented in this study are available from the Longitudinal Aging Study Amsterdam (LASA) and may be requested for research purposes. More information on data requests can be found on: www.lasa-vu.nl.

## Contributors

EH conducted the statistical analyses and drafted the manuscript. All authors critically revised drafts of the manuscript, approved the final version and agreed to be accountable for all aspects of the work.

## Patient consent for publication

Not required.

## Ethics approval

This study is conducted in line with the Declaration of Helsinki and received approval by the medical ethics committee of the VU University medical centre.

## Data availability statement

The datasets generated during the current study are not publicly available, but the data underlying the results presented in this study are available from the Longitudinal Aging Study Amsterdam (LASA) and may be requested for research purposes. More information on data requests can be found on: www.lasa-vu.nl.

